# USING SYSTEM DYNAMICS MODELING TO ASSESS THE IMPACT OF CONNECTICUT’S GOOD SAMARITAN LAWS: A PROTOCOL PAPER

**DOI:** 10.1101/2022.01.06.22268677

**Authors:** Syed Shayan Ali, Nasim S. Sabounchi, Robert Heimer, Gail D’Onofrio, Colleen Violette, Katherine LaWall, Rebekah Heckmann

**Affiliations:** Department of Emergency Medicine, Yale School of Medicine, New Haven, CT, USA; Department of Health Policy and Management, City University of New York (CUNY) Graduate School of Public Health and Health Policy, New York, NY, USA; Department of Epidemiology, Yale School of Public Health, New Haven, CT, USA; Connecticut Department of Public Health, Hartford, CT, USA

**Keywords:** System dynamics modeling, group model building, opioid overdose deaths, opioid use disorder, Good Samaritan laws

## Abstract

**Background:** We applied a participatory system dynamics (SD) modeling approach to evaluate the effectiveness and impact of Connecticut’s Good Samaritan Laws (GSLs) that are designed to promote bystander intervention during an opioid overdose event and reduce opioid overdose-related adverse outcomes. Our SD model can be used to predict whether additional revisions of the statutes might make GSLs more effective. SD modeling is a novel approach for assessing the impact of GSLs; and, in this protocol paper, we describe its applicability to our policy question, as well as expected outcomes of this approach.

**Methods:** This project began in February 2021 and is expected to conclude by March 2022. During this time, a total of six group model-building (GMB) sessions will have been held with key stakeholders to elicit feedback that will, in turn, contribute to the development of a more robust SD model. Session participants include bystanders who witness an overdose, law enforcement personnel, first responders, pharmacists, physicians, and other health care professionals who work in at least two major metropolitan areas of Connecticut (New Haven and Hartford). Due to the restrictions imposed by the COVID-19 pandemic, the sessions are being held virtually via Zoom. The information obtained during these sessions will be integrated with a draft SD model that has already been developed by the modeling team as part of a previous CDC-funded project. Model calibration and policy simulations will then be performed to assess the impact of the current GSLs and to make recommendations for future public policy changes.

**Discussion:** An SD modeling approach enables capture of complex interrelationships among multiple health outcomes to better assess the drivers of the opioid epidemic in Connecticut. The model’s simulation results are expected not only to align with current real-world data but also to recreate historical trends and infer future trends in a situationally relevant fashion. This will facilitate the work of policy makers who are devising and implementing time-sensitive changes to address opioid overdose-related deaths at the state level. Replicating our approach as described can be applied to make similar improvements in other jurisdictions.

**CONTRIBUTIONS TO THE LITERATURE:**

- System dynamics (SD) modeling and group model-building (GMB) approaches enable the group to start with a simple concept model and apply the collective knowledge of the group to finish the session with a much more developed model that can produce impressively accurate simulation results.
- The model will be used to understand the impact of Connecticut’s Good Samaritan Laws (GSLs), as well as their limitations, and to deduce factors to further improve public health laws to counter opioid overdose-related deaths.
- The approach can be applied to other jurisdictions, taking into account local conditions and existing Good Samaritan legislation.

## BACKGROUND

In late 2017, after the deaths due to opioid-related overdoses had almost tripled within a decade (1), the US Department of Health and Human Services declared opioid use-related casualties to be a national public health emergency and called for immediate augmentation of interventions. Deaths resulting from opioid-related overdoses are now the leading cause of preventable human deaths in the US (2, 3). In Connecticut, drug overdose fatality rates showed a significant increase for all races combined, estimated to be 3.6 deaths per 100,000 population per year, making it an exemplar of the opioid overdose trend in the New England region of our country (4).

The persistent increase in opioid poisoning deaths remains a national public health crisis, demanding innovative policy interventions to counter this crisis. One major intervention is the widespread distribution of naloxone, an opioid antagonist that reverses the adverse effects of opioids. The intervention has targeted people who misuse opioids and their families through expanded prescribing and community distribution (5–9). Almost all states have enacted some form of a Good Samaritan law (GSL) to reduce deaths from opioid overdose. These GSLs are intended to provide legal protection against liability and arrest for bystanders who give assistance during an overdose incident by administering naloxone. In addition, they can protect first responders and individuals who prescribe or provide naloxone that is administered during an episode. Connecticut’s GSL was originally passed in 2011 and has been updated and expanded several times since 2014 (10, 11).

Previous efforts to curb the opioid overdose epidemic have focused on restricting the supply of prescription narcotics used for analgesia; however, the epidemic has transitioned from being driven by pharmaceutical to non-pharmaceutical opioids. Interventions that merely restrict the supply of pharmaceuticals has proven inadequate to reduce opioid overdose related mortality and morbidity. The changing nature of the epidemic requires novel interventions based on harm reduction and demand reduction, and these are crucial now more than ever as the COVID pandemic has accelerated the incidence of opioid overdoses. Without implementation of effective public health interventions, the opioid crisis is expected to persist in the next decade due to several factors, including an expected increase in the number of individuals using illicit opioids, easy availability of illicit opioids, and the increase in lethality of these illicit opioids due to the highly potent synthetic opioid fentanyl that now dominates the US supply (12, 13).

Responding to the opioid epidemic is just one of many public health issues that present complex challenges requiring a systems approach to assess the different components of health and prevention systems. Novel approaches are needed to explore component interactions and devise policy options that consist of the most effective and efficient arrangements of multiple resources available within a system. One such approach is system dynamics (SD) modeling which, according to Homer and Hirsch, involves “an interlocking set of differential and algebraic equations that are devised from a broad spectrum of measured and experiential data” (14).

We are employing an SD approach to assess the numerous factors that impact the influence of the GSLs in Connecticut and will incorporate measurement of multiple factors and their concurrent variance to determine this influence. The statewide modeling will focus on the following three primary health outcomes: (1) emergency department visits for drug misuse and overdose, (2) behavioral changes of bystanders, and (3) overdose deaths. We believe that this model will prove to be superior to the use of more traditional methods such as surveys, participant interviews, and state level comparisons of policy modifications in assessing the effectiveness of the GSL. By providing insights that cannot be obtained through these other means of analysis, we expect that the modeling outputs will provide a more structured understanding of the effectiveness of the GSLs by explaining feedback loops and endogenous sources of system behavior (15).

The Centers for Disease Control’s (CDC) Prescription Drug Overdose (PDO) Prevention for States (PfS) program was designed to promote and evaluate state-level interventions for “preventing drug overuse, misuse, abuse, and overdose;” and this policy evaluation represents this group’s attempt to address the goal articulated by the CDC. In response to recommendations from the National Academies of Sciences, Engineering and Medicine (NASEM), the Federal Drug and Food administration (FDA) has now accepted an SD modeling approach as their nationwide approach to assess the impact of GSLs that have been implemented throughout the country (16–19)

We developed this protocol paper as means of describing SD modeling as a novel way of assessing the impact of public health interventions and developing and testing effective public health policies that address complex policy problems.

## METHODS/DESIGN

### Study Setting

Connecticut has witnessed a steady increase in total overdose deaths among residents, from 357 deaths in 2012 to 1,374 deaths in 2020, an increase of almost 285% (20). Almost half of these deaths occurred at a residence. In 2016, the state ranked 11^th^ in terms of opioid overdose related deaths in the country with 27.4 deaths per every 100,000 people (21). Many of Connecticut’s neighboring states are also among the top 10 states for their rates of overdose deaths. This includes New Hampshire (39 deaths/100,000 people), Massachusetts (33 deaths/100,000 people), Rhode Island (30.8 deaths/100,000 people), and Maine (28.7 deaths/100,000 people)(22). Regardless of gender, race, and age, the opioid epidemic continues to worsen in Connecticut (22). Males have higher rates of overdose deaths than females. These demographic trends are consistent throughout most of the nation. The average age of death varies significantly, with whites dying at a younger age than blacks and Hispanics each year in Connecticut (20). Most overdose deaths in CT involved illicit drugs, such as fentanyl, heroin, and cocaine, rather than prescription drugs (4). Heroin was the most common substance leading to opioid-related deaths among CT residents from 2012-2017 (22, 23). More recently, fentanyl has predominated and now is involved in 85-90% of all accidental drug poisonings (20).

The passage of the CT GSL in 2011 was intended to reduce the reluctance to administer naloxone by providing immunity, by statute, from criminal prosecution to the bystander who responded to an overdose even if law enforcement found evidence of illicit drug use or presence at the scene. It expanded the statute that already allowed licensed health care professionals to administer an opioid antagonist to a drug user without fear of liability for civil damages or criminal prosecution. The scope of the GSL was expanded in 2012 to offer protection from civil penalties to prescribing practitioners, pharmacists, law enforcement agencies, emergency medical service providers, government agencies, community health organizations and other entities that distribute naloxone for use in responding to opioid overdoses. It was expanded again in 2014 to protect against liability for civil damages or criminal prosecution for the possession of drugs or related paraphernalia to any person who administered an opioid antagonist during a suspected overdose. A related statute passed in 2015 that allowed pharmacists to sell naloxone to anyone requesting it by creating a statewide standing order, in effect turning a prescription medication into one available over the counter (11).

### Conceptual framework

Group model-building (GMB) is a form of participatory SD modeling with which we are capturing the knowledge and perceptions of stakeholders and domain experts. These will be analyzed and integrated to assess the effectiveness and impact of the GSLs on bystander behavior in reducing opioid-related deaths in Connecticut. The purpose of a GMB approach is twofold – to use participants’ understanding of a complex problem to reach common ground when designing policy innovations and to gain support from participants for implementation of model-based policy recommendations (24–26). We facilitate the GMB sessions using scripts that have been developed, revised, and validated by various SD expert modelers through a collaborative effort. The scripts are documented and made accessible on the Scriptapedia platform to make GMB in SD more science than art (27–29). The GMB approach engages stakeholders in a process that yields compelling, graphical outputs that inform both challenges of and solutions to complex systems problems. During the workshop, participants are oriented to systems thinking and modeling and use the causal-loop diagramming (CLD) approach to describe interdependencies and sketch feedback structures.

SD modeling and GMB have recently been applied to implementation science in complex public health problems. For example, SD has been applied to improving developmental-behavioural screening and detection of related disorders in primary care settings (30). SD has also been a useful tool for understanding the complexity and dynamics around the adaptation and implementation of financial policies to increase health insurance in Mexico and reduce financial inequities (31). Also, participatory SD modeling using the GMB approach has been used for improving evidence-based practice for PTSD patients in the VA (32, 33). Another study has used SD modeling to study the long-term consequences of formal guidelines around mammography screening and how these are implemented, interpreted, and acted upon by various stakeholders (34).

There remains a gap in adopting SD and participatory modeling approaches for real-time public policy analysis and assessing the impact of implementing identified policy changes at the state level. In facilitating the GMB sessions for this project that evaluates the GSLs, we are engaging key stakeholders to introduce perspectives from a broad range of expertise and experiences such that the conceptual model, with its system structures and feedback loops underlying the opioid crisis in Connecticut, has the potential for identifying policy levers for improving system outcomes.

### Study design

Our study sets out to measure multiple factors and their simultaneous variance to determine the effectiveness of the GSLs in Connecticut. Model calibration will be used to make estimates for the parameter values that were not readily available in literature. A maximum likelihood estimation approach will be used to create simulations which align best with real-world data, replicating historical trends and creating estimations for the future. To accomplish these aims, we have planned a four-part SD modeling approach. First, we have established a baseline data framework from publicly available data and created a preliminary model to develop questions to better understand how the multiple factors interact. Second, the questions are then posed to stakeholders during the GMB for directed data collection on the nature of the interactions. Finally, we will interpret the data to refine the SD model and assess if the refined model has improved predictive value.

#### Preparing for GMB

Baseline data to calibrate the model are obtained from several sources. Data on nonfatal opioid-involved overdoses come from emergency medical reporting and the Connecticut Hospital Association (CHIME) discharge records. Data on fatal opioid-involved overdoses come from the CT Office of the Chief Medical Examiner. Data on illicit drug use and related arrests come from several sources compiled by Substance Abuse and Mental Health Services Administration (SAMHSA) reports from 2015 to 2020. Data on the rate of opioid prescriptions will be retrieved from the CDC report on U.S. state prescribing rates. Initial information on knowledge of the GSL and fear of calling 911 to report an overdose come from a study conducted by the Connecticut Department of Public Health (CT DPH) and Central Connecticut State University (CCSU) and a High Intensity Drug Trafficking Area’s (HIDTA) Heroin Response Strategy project (35, 36). Once the baseline data had been compiled, they were used to build a preliminary SD model that identified putative interactions between events that occur to individuals within opioid using populations. The interactions were also used to formulate questions for the GMB sessions that are incorporated in scripts that allow all GMB sessions to cover the same issues (28).

#### Conducting Group Model Building Sessions

In practice, each GMB session is two hours long and conducted by trained meeting *facilitators, modelers*, and *recorders*. The *lead facilitator* initiates the meeting, provides a brief introduction to the project, the *modeling team*, and *the participants*, and reviews the project’s goals and intention. Following this introduction, *the participants* are divided into small groups led by *facilitators* who guide *the participants* through scripts to facilitate the discussions around defining key factors. The *participants* then construct, based on their insider knowledge, behavior-over-time graph (BOTG) diagrams that focus on trends central to the problem definition and that serve to refine the modeling process (25). Participants have 15 minutes to draw the graphs with prompting from *facilitators* if needed to highlight graphically their identification of a problem related to the opioid epidemic and its expected trajectory over time. Throughout the session, the *recorder* is entering a verbal account of the session into a Microsoft Word document to identify recurrent themes and key insights. The *facilitator* then reviews the graphs to form clusters and themes.

The *modeler* uses the graphs to identify a few key variables named by participants, as well as the causal links between variables to begin forming a causal loop diagram (CLD). A CLD is a directed graph that depicts how different variables in a system are interrelated. The CLDs consist of a set of variables and positive and negative causal links. Causal links represent a relation between the two variables in the form of cause and effect. Relations between variables form an abstract diagram that consists of few loops, which can depict complicated interactions about a particular problem or issue. As shown in the accompanying diagram (Figure 1), two distinct kinds of causal loops exist. *<u>Reinforcing loops</u>* produce exponential increase or decrease. Reinforcing loops are identifiable by an *even* number of negative causal links in them. *<u>Balancing loops c</u>*ounter change in an attempt to bring things to a stable state and keep them there and consist of an *odd* number of negative causal links. As an example, a reinforcing loop describes greater availability of Narcan due to increasing acceptability of Narcan after lives are saved because of its administration (indicated by R1 in Figure 1), hence reducing overdose mortality; while in a balancing loop, an increase in the number of overdose incidents leads to more overdose fatalities and, initially, to a reduced number of people with opioid use disorder. Throughout the sessions, we are continuing to make note of these interrelationships to focus on possible interventions and modifications to devise feasible and acceptable policy changes that might contribute to our continued efforts to curb the opioid use crisis in Connecticut

**Figure 1:**
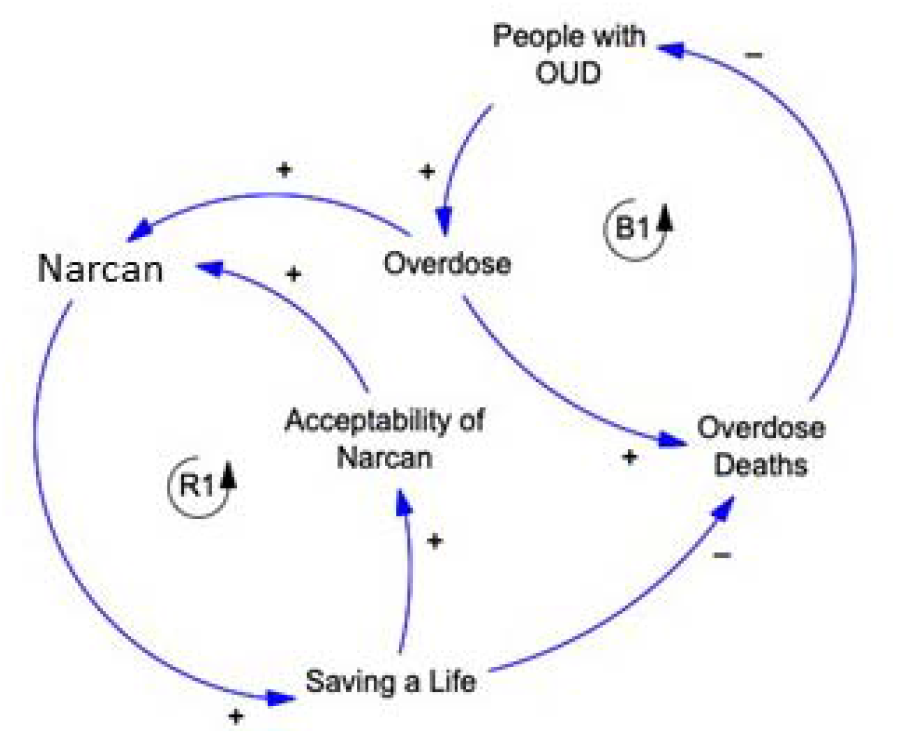
R1 depicts a reinforcing loop as Narcan use leads to lives saved, ultimately leading to more acceptance of Narcan use; whereas, B1 depicts a balancing loop that shows that overdose deaths initially lead to a reduction in the number of people with opioid use disorder.

As more complicated CLDs emerge from the GMB, they will still contain just a few basic elements: the variables, the links between them, the signs on the links (which show how the variables are interconnected), delays, and the sign of the loop. By representing a problem or issue from a causal perspective, both modelers and participants become more aware of the structural forces that produce puzzling behavior. The resulting diagrams also provide a visual representation that can be used to communicate that understanding with others.

The *facilitator* concludes the session by asking the teams to consider insights they gained from the exercise and build their own CLDs, explaining factors driving overdose deaths by adding other variables and drawing arrows connecting them to the existing loops. During this step, each small group has the opportunity to draw attention to more important loops and comment on the relative influence of one loop on the other loops in the diagram. By using CLDs to create stories about complex issues, the group of stakeholders can make more explicit the modelers’ understanding of the interrelationships within a system’s structure. To ensure that as much information as possible is gleaned from these exercises, the *modeling team* engages in a reflection session led by the *debriefer* immediately following the session.

For this project, six GMB sessions will be conducted in Connecticut over a 15-month period that ends in March 2022. While the initial plan was to focus on participants from just two parts of the state, the ongoing COVID-19 pandemic forced the GMB sessions to occur virtually and, thereby, facilitated broader statewide representation. Over the course of the study, the sessions are enabling the modeling team to capture, understand, and portray the complex interrelationships among several key health outcome measures that are impacted by the opioid epidemic in Connecticut. These outcomes include ED visits due to overdose, behavioral changes of bystanders, changes in perception of the risk of drug use, awareness of harm reduction policies, and overdose deaths, Additionally, the sessions have helped the project team to acquire a deeper understanding of the interrelationships among key stakeholders. To maximize understanding of the efficacy of current public health laws, participants are being selected from key stakeholder groups, including pharmacists, other healthcare providers, emergency medical service clinicians, law enforcement personnel, individuals who have experienced or witnessed an overdose, family members of individuals who have experienced overdose, fire-fighting personnel, and other first responders. Participants are chosen carefully to provide adequate representation of the stakeholder groups that play a key role in responding to the opioid emergency. A particular emphasis has been made to include bystanders who directly benefit from the GSL, law enforcement agents who have to abide by the current laws which may differ from previous practices, and healthcare professionals and pharmacists who benefit from the protection in amendments added after 2011. The sessions have engaged participants in exercises of narrative elicitation, computerized causal loop diagraming of the underlying problems, and construction of behavior over time graphs. Stakeholder participation exercises and scripts are designed to focus attention and elicit information regarding the implementation and effectiveness of Good Samaritan Laws, with a specific focus on encouraging bystander intervention during an opioid overdose event. But participants are also encouraged to generate CLDs that identify the factors that, in their experience, influence the willingness of bystanders to act.

Existing data narratives presented in the form of a CLD serve as the means of obtaining input and to prime the group discussion. With help from participants and the behavior-over-time graphs that they produce, the variables and the identified relationships in the CLD are updated throughout the sessions and confirmed with the participants to minimize misinterpretation. Input from each session is incorporated to enhance the next session with new information to confirm or modify the relationship. Throughout the sessions, the *recorder* continues to make notes of these relationships and possible interventions and modifications to help improve the SD model by devising feasible and acceptable policy changes that might contribute to efforts to curb the opioid use crisis in Connecticut.

Immediately following each session, the *facilitators*, in collaboration with the *recorders*, analyze how the GMB session improved participant understanding of systems thinking. The team also identifies additional information and participant insights that can be used to improve the specificity and sensitivity of the preliminary SD model, in addition to studying how each GMB session participant perceived the usefulness of the session for understanding the policy’s intent and effect. The modeling team administers a GMB knowledge assessment tool to capture the level of subject knowledge of each participant using a structured set of questions (33). This allows modelers to weigh the CLDs based on their interpretation of the level of knowledge participants have brought to the CLD exercises.

At the time this manuscript was developed, three sessions had been completed. These initial sessions involved the two main activities described above: constructing behavior-over-time graphs and contributing to causal loop diagramming. Each activity was initially performed by sub-groups and concluded with the full group engaging in discussion. For the BOTG activity, we asked participant to answer the following question graphically: “What are the facilitators and barriers that impact the effectiveness of the Good Samaritan Laws on distributing naloxone and reducing opioid-related deaths in Connecticut?” In some cases, some prompting was needed to help initiate the process. The participants were then asked to rank their graphs over time with the most important/favorite factors as first and the least important/least favorite factors as last. They were asked to either display their graphs on their camera using the Zoom application or to draw the graph on the white board tab of the software. They were then asked to describe briefly what they drew to engage in discussion. In the next activity, the participants were shown a basic CLD and engaged in forming additional loops to expand upon the relationships between various factors.

From the sessions that we have conducted thus far, we have learned that longer sessions (approximately 2 hours) with fewer participants (5-10 participants) in each session allow for more engaging and thought-provoking sessions that seem to elicit more detailed feedback than sessions with more participants. For the subsequent three sessions that are to follow, we plan to build upon the data that has been collected in the first three sessions. We are working to derive main take-away lessons – persistent themes, opportunities to intervene, barriers to implementing potentially effective interventions – from the first three sessions and present them in CLDs to participants to engage in discussion regarding policy planning. The sessions will involve review of balancing and reinforcing loops and a review of potential policy changes.

#### Analysis plan

The results of the all six modeling workshops and their follow-up assessments will then be integrated to refine the preliminary SD model that had been developed by the modeling team as part of a previous CDC-funded project to understand how laws and policies can be more effectively implemented to control the epidemic (37). We will be using tools and notation from established systems science analytic methods to help us organize the feedback that we receive during these sessions (38). These tools will allow the team to move the focus from a linear cause-and-effect paradigm to a model that includes feedback loops and interlocking causes and effects. Additionally, the analysis of the information collected during the sessions will help the project team acquire a deeper understanding of the interrelationships among key stakeholders. This understanding is key to developing actionable interventions that have both a high likelihood of reducing opioid overdose mortality and being acceptable and implementable by a broad swath of stakeholders and the affected population. The team will develop policy improvement recommendations integrating the information obtained from the SD modeling exercises performed during GMB sessions and GMB post-session knowledge assessments using computerized SD model simulations with time varying elements. The model will incorporate available state agency data including the number of EMS episodes and ED visits for opioid drug overdose, the number of opioid-involved overdose deaths, and use of treatment services for opioid use disorder. Data from the GMB sessions will help put bounds on unknowns such as the number of people misusing prescription medications and street drugs, changes in drug markets and drug testing for the presence of fentanyl, the behavioral changes of bystanders, and the number of police officers and members of the public who have GSL knowledge. The output will be an upgraded SD model. The model will then be used to construct “what-if” analyses to determine and rank in priority the constituent parts of the system and identify the policy innovations that are likely to result in the largest change in outcome measured as a reduction in opioid overdose deaths.

## DISCUSSION

SD modeling enables us to capture and analyze the complex interrelationships that exist between multiple public health practices that ultimately impact the opioid epidemic in Connecticut. This modeling approach allows researchers to investigate the delayed and long-term impacts of policy changes such as the GSLs by facilitating construction of a simulation framework. By effectively replicating historical trends, SD modeling estimates future effectiveness of interventions without requiring extensive involvement of study participants or the need for costly pilot studies prior to widespread implementation of policy changes.

In our current effort, we have focused on GSLs that were passed and progressively expanded in Connecticut beginning 10 years ago. SD modeling can prove beneficial as an analytic tool to assess the expected impact of modifications which, otherwise, might not demonstrate observable outcomes for several years, as policy changes often take time before widespread adoption. Modeling can also help to integrate data sets by producing data-driven simulations that can account for missing or incomplete variables where the basic direction of causality between important variables is thought to be understood. The GMB sessions focus attention on directionality. Additionally, this type of modeling can be especially helpful when the definition of the system of interest (in our case, the state of Connecticut) is clear.

We employed the SD modelling approach to assess the effectiveness of the current CT GSL; however, we intend to use the model to assess other strategies to combat the opioid crisis. Our model structure and feedback loops are very relevant to the opioid epidemic and, thus, can be easily calibrated to apply to different locations for the same purpose. A key benefit of the GMB sessions is that participating stakeholders come to understand the model and have bought into the process of using models to predict the impact of policy changes. Thus, one of the biggest advantages of using the SD model is allowing public health decision makers in Connecticut to appreciate, employ, and even update the current SD model and anticipate the impact of new or evolving policies that may come their way. An additional benefit of the project, as a whole, is the development of an approach that can be applied elsewhere even if circumstances specific to Connecticut may differ. The GMB sessions are central to the process of making a general approach that can be attuned to local conditions.

### Limitations and future recommendations

Due to limitations imposed by the COVID-19 pandemic, sessions are conducted virtually via Zoom. This initially created technical difficulties for some of our participants and interrupted optimal interactions between the *facilitators, modeler*, and *participants*. However, this challenge led the project team to develop a set of best practices for virtual GMB facilitation that will likely become increasingly useful as virtual research becomes a more mainstream method for this type of evaluation.

## Supporting information

Checklist

## Data Availability

All data produced in the present study are available upon reasonable request to the authors

## LIST OF ABBREVIATIONS

SD: Systems dynamic modelling
GMB: Group-based model-building
CLD: Causal Loop Diagrams
GSL: good Samaritan law

## DECLARATION

### Ethics approval and consent to participate

This project is a collaborative work of researchers from Yale University and City University of New York (CUNY). The project was approved by Yale University IRB on 10/6/20 (Yale IRB #2000028908) and CUNY IRB on 12/30/20 (CUNY IRB number: HRPP #2020-0958).

### Consent for publication

Not applicable

### Availability of data and materials

Not applicable

### Authors’ contributions

SSA, NS, RH, GD, RH drafted the manuscript and contributed to the design of the study. CV and KL collaborated with the modeling team to help develop the project’s specifications and provide guidance on Connecticut Department of Public Health policies for this public health initiative. All authors read and approved the final manuscript.

## Acknowledgements

We would like to acknowledge Susan Logan from the Connecticut Department of Public Health for her support of the project.

## Authors’ information

(optional)

## REFERENCES

1. Scholl L, Seth P, Kariisa M, Wilson N, Baldwin G. Drug and opioid-involved overdose deaths— United States, 2013–2017. Morbidity and Mortality Weekly Report. 2019;67(5152):1419.

2. Watson DP, Ray B, Robison L, Huynh P, Sightes E, Brucker K, et al. Lay responder naloxone access and Good Samaritan law compliance: postcard survey results from 20 Indiana counties. Harm reduction journal. 2018;15(1):18.

3. Chen Q, Larochelle MR, Weaver DT, Lietz AP, Mueller PP, Mercaldo S, et al. Prevention of prescription opioid misuse and projected overdose deaths in the United States. JAMA network open. 2019;2(2):e187621–e.

4. Wu ZH, Yong Q, Walker JM, Grady JJ, Laurencin CT. Fentanyl, Heroin, and Cocaine Overdose Fatalities are Shifting to the Black Community: An Analysis of the State of Connecticut. J Racial Ethn Health Disparities. 2021.

5. Albert S, Brason FW, 2nd, Sanford CK, Dasgupta N, Graham J, Lovette B. Project Lazarus: community-based overdose prevention in rural North Carolina. Pain Med. 2011;12 Suppl 2:S77–85.

6. Bennett AS, Elliott L. Naloxone’s role in the national opioid crisis-past struggles, current efforts, and future opportunities. Transl Res. 2021;234:43–57.

7. Doe-Simkins M, Walley AY, Epstein A, Moyer P. Saved by the nose: bystander-administered intranasal naloxone hydrochloride for opioid overdose. Am J Public Health. 2009;99(5):788–91.

8. Keane C, Egan JE, Hawk M. Effects of naloxone distribution to likely bystanders: Results of an agent-based model. Int J Drug Policy. 2018;55:61–9.

9. Seal KH, Thawley R, Gee L, Bamberger J, Kral AH, Ciccarone D, et al. Naloxone distribution and cardiopulmonary resuscitation training for injection drug users to prevent heroin overdose death: a pilot intervention study. J Urban Health. 2005;82(2):303–11.

10. DMHAS. Opioid Overdose Prevention/Naloxone (Narcan) Initiative. Connecticut Department of Mental Health & Addiction Services; 2019.

11. The Good Samaritan Law in CT. Available from: https://www.drugfreect.org/resources/ct-good-samaritan-law/.

12. Cicero TJ, Ellis MS, Kasper ZA. Increased use of heroin as an initiating opioid of abuse. Addict Behav. 2017;74:63–6.

13. Wilson N KM, Seth P, Smith H IV, Davis NL.. Drug and Opioid-Involved Overdose Deaths — United States, 2017–2018. MMWR Morb Mortal Wkly Rep 2020;69:290–297. DOI.

14. Homer JB, Hirsch GB. System dynamics modeling for public health: background and opportunities. American journal of public health. 2006;96(3):452–8.

15. Atkinson J-A, Wells R, Page A, Dominello A, Haines M, Wilson A. Applications of system dynamics modelling to support health policy. Public Health Res Pract. 2015;25(3):e2531531.

16. Naumann RB, Austin AE, Sheble L, Lich KH. System dynamics applications to injury and violence prevention: a systematic review. Curr Epidemiol Rep. 2019;6:248–62.

17. Wakeland W, Nielsen A, Geissert P. Dynamic model of nonmedical opioid use trajectories and potential policy interventions. Am J Drug Alcohol Abuse. 2015;41(6):508–18.

18. Wakeland W, Schmidt T, Gilson AM, Haddox JD, Webster LR. System dynamics modeling as a potentially useful tool in analyzing mitigation strategies to reduce overdose deaths associated with pharmaceutical opioid treatment of chronic pain. Pain Med. 2011;12 Suppl 2:S49–58.

19. FDA Opioid Systems Modeling Effort. Available from: https://www.fda.gov/drugs/information-drug-class/fda-opioid-systems-modeling-effort.

20. Opioids and Prescription Drug Overdose Prevention. Connecticut Department of Public Health. Unintentional drug overdose deaths in Connecticut: a fact sheet – 2020. 2020.

21. Opioids and Prescription Drug Overdose Prevention. Available from: https://portal.ct.gov/dph/Health-Education-Management--Surveillance/The-Office-of-Injury-Prevention/Opioids-and-Prescription-Drug-Overdose-Prevention-Program.

22. Ankrah J. Connecticut’s Opioid Epidemic: A Glimpse of the Past Five Years. DataHaven, 2018.

23. 2012-2017 Accidental Drug-Related Deaths. Connecticut Office of the Chief Medical Examiner : https://data.ct.gov/Health-and-Human-Services/Accidental-Drug-Related-Deaths-2012-2017/ecj5-r2i9.

24. Ja V. Group model-building: tackling messy problems. System Dynamics Review. 1999.

25. Andersen DF, Richardson GP. Scripts for group model building. System Dynamics Review. 1997;13:107–29.

26. Andersen DF, Richardson GP, Vennix JAM. Group model building: adding more science to the craft. System Dynamics Review. 1997;13(2):187–201.

27. Hovmand PS. Group Model Building and Community-Based System Dynamics Process. Community Based System Dynamics. New York, NY: Springer New York; 2014. p. 17–30.

28. Hovmand PS, Andersen DF, Rouwette E, Richardson GP, Rux K, Calhoun A. Group Model-Building ‘Scripts’ as a Collaborative Planning Tool. Systems Research and Behavioral Science. 2012;29(2):179–93.

29. WikiBooks. Available from: https://en.wikibooks.org/wiki/Scriptapedia.

30. Sheldrick RC, Breuer DJ, Hassan R, Chan K, Polk DE, Benneyan J. A system dynamics model of clinical decision thresholds for the detection of developmental-behavioral disorders. Implementation Science. 2016;11(1):1–14.

31. Nigenda G, González-Robledo LM, Juárez-Ramírez C, Adam T. Understanding the dynamics of the Seguro Popular de Salud policy implementation in Mexico from a complex adaptive systems perspective. Implementation Science. 2015;11(1):1–12.

32. Lounsbury D, Kibbe D, Rollins J, Zimmerman L, editors. From blank page to local optimization: participatory systems modeling to improve local evidence based practice implementation. Implementation Science; 2020: Bmc Campus, 4 Crinan St, London N1 9xw, England.

33. Zimmerman L, Lounsbury DW, Rosen CS, Kimerling R, Trafton JA, Lindley SE. Participatory System Dynamics Modeling: Increasing Stakeholder Engagement and Precision to Improve Implementation Planning in Systems. Administration and Policy in Mental Health and Mental Health Services Research. 2016;43(6):834–49.

34. Karanfil O, Sterman JD. A dynamic model for health screening: misperceptions, feedback and long term trends in screening mammography. Implementation Science. 2015;10(1):A50.

35. HIDTA. Heroin Response Strategy - 911 Good Samaritan Law Cornerstone Project - Preliminary State Report Connecticut: High Intensity Drug Trafficking Areas (HIDTA) Program - White House Office of National Drug Control Policy; 2019.

36. Hawk KF, Doernberg M, D’Onofrio G, Heimer R, Diaz-Matos LF, Jenkins M, et al. Knowledge of Connecticut’s Good Samaritan Law Among Connecticut Residents Who Have Witnessed an Overdose. 2019.

37. Sabounchi NS, Heckmann R, D’Onofrio G, Walker J, Heimer R. Assessing the Impact of the Good Samaritan Law in the State of Connecticut: A System Dynamics Approach. Health Research Policy and Systems. Accepted December 2021.

38. Sterman J. System Dynamics: systems thinking and modeling for a complex world 2002.

