## Supplementary material for "USING SYSTEM DYNAMICS MODELING TO ASSESS THE IMPACT OF CONNECTICUT’S GOOD SAMARITAN LAWS: A PROTOCOL PAPER": Checklist

### TIDieR-PHP Checklist: a reporting guideline for population health and policy interventions

The citation for the TIDieR-PHP explanation and elaboration article is: Campbell M, Katikireddi SV, Hoffmann T, Armstrong R, Waters E and Craig P. TIDieR-PHP: a reporting guideline for population health and policy interventions, explanation and elaboration. BMJ 2018;360:k1079. http://dx.doi.org/10.1136/bmj.k1079

| TIDieR-PHP is intended to complement and be used as an extension to the appropriate reporting guideline for the study design being used (e.g. CONSORT, SPIRIT, STROBE, or TREND) | | | |
|---|---|---|---|
| **TIDieR-PHP item** | **Item description** | **Page in manuscript where item is reported** | **Other*** |
| 1 Brief name | Provide the name or a phrase that describes the intervention | 7 | |
| 2 Why | Describe the logic, mechanisms or rationale of the intervention clearly linking intervention elements to the expected effect on immediate or longer term outcomes (or both) | 7 | |
| 3 What - materials | Describe any materials used in the intervention (including any online appendices or URL for further details). For example:<br>• informational materials (may include those provided to recipients of the intervention or in training of intervention providers)<br>• nature and value of any benefit provided (e.g. cash, voucher, meal)<br>• any physical resources or infrastructure provided as part of the intervention | 7 | |
| 4 What and How | Describe how the intervention was planned, established, and intended to be delivered. Depending on the type of intervention, it may be useful to consider:<br>• how sources of funding for the intervention and the service providers were obtained, how users were enrolled and the service delivered<br>• how any payments were made or benefits delivered, how qualifying conditions were implemented<br>• the entity being regulated, the scope of the regulation, permitted level of use; procedures for monitoring or enforcing compliance, and any sanctions for non-compliance<br>• how people were exposed to the intervention, whether it was provided to individuals or larger populations<br>• any underpinning legislation including name, date passed and legislative body | 6 | |

### TIDieR-PHP Checklist: a reporting guideline for population health and policy interventions

| TIDieR-PHP item | Item description | Page in manuscript where item is reported | Other* |
|---|---|---|---|
| 5 Who provided | Describe the provider of the intervention, including legal status and powers, field organisations and staff responsible for planning, implementation, monitoring and enforcement. Where relevant, describe intervention provider expertise and training (general or specific to the intervention) | 7-19 | |
| 6 Where | Describe the type of location (e.g. school, community centre) and the geographical scope of the intervention (e.g. national, regional, city-wide). Where relevant, describe the historical, cultural, socioeconomic, or political background to the intervention | 6 | |
| 7 When and how often | Describe when the intervention was implemented, how long it remained in place, and if applicable, the number, duration and scheduling of occasions | 6 | |
| 8.1 Planned variation | Describe and provide the reason for any variation or tailoring that was planned or allowed for in the design of the intervention. Examples include differences between locations, geographical areas, population subgroups or over time | 10 | |
| 8.2 Unplanned variation | Describe and provide the reason for any unplanned variation or modifications in the intervention (e.g. between different locations, geographical areas, population subgroups, or over time) that were made after the intervention commenced | N/A | |
| 9.1 How well | Describe any strategies used or actions taken to maintain fidelity of the intervention (i.e., to ensure that the intervention was delivered as intended) | 14-15 | |
| 9.2 How well - delivery | Describe the fidelity of the intervention (i.e., the extent to which the intervention was delivered as intended) | 14-15 | |

*If the information is not provided in the primary paper, give details of where this information is available (eg, protocol, other published papers (provide citation details), or a website (provide the URL))
